# Patients with Asthma and Chronic Obstructive Pulmonary Disease (COPD) have increased levels of plasma inflammatory mediators upregulated in severe COVID-19

**DOI:** 10.1101/2021.01.23.21250370

**Authors:** Nathalie Acevedo, Jose Miguel Escamilla-Gil, Héctor Espinoza, Ronald Regino, Jonathan Ramírez, Lucila Florez de Arco, Rodolfo Dennis, Carlos Torres-Duque, Luis Caraballo

## Abstract

**Background:** Chronic obstructive pulmonary disease (COPD) is associated with increased risk of severe COVID-19, but the mechanisms are unclear. Besides, patients with severe COVID-19 have been reported to have increased levels of several immune mediators.

**Objective:** To perform an immunoproteomic profiling of dysregulated plasma proteins in patients with asthma and COPD and to evaluate their relationship with biomarkers of severe COVID-19.

**Methods:** Ninety-two proteins were quantified in 315 plasma samples from adult subjects (age 40-90 years) including 118 asthmatics, 99 COPD patients and 98 healthy controls, that have been recruited in two reference pneumology clinics in Colombia before the beginning of the COVID-19 pandemic. Protein levels were compared between each disease group and healthy controls.

Significant proteins were compared to the gene signatures of SARS-CoV-2 infection reported in the “COVID-19 Drug and Gene Set Library” and with known protein biomarkers of severe COVID-19.

**Results:** Forty-one plasma proteins showed differences between patients and controls. Asthmatic patients have increased levels in IL-6 while COPD patients have a broader systemic inflammatory dysregulation driven by HGF, OPG, and several chemokines (CXCL9, CXCL10, CXCL11, CX3CL1, CXCL1, MCP-3, MCP-4, CCL3, CCL4 and CCL11). These proteins are involved in chemokine signaling pathways related with response to viral infections and some, were found up-regulated upon SARS-CoV-2 experimental infection of Calu-3 cells as reported in the COVID-19 Related Gene Sets database. An increase of HPG, CXCL9, CXCL10, IL-6, MCP-3, TNF and EN-RAGE has also been found in patients with severe COVID-19.

**Conclusions:** COPD patients have altered levels of plasma proteins that have been reported increased in patients with severe COVID-19. Our study suggests that COPD patients have a systemic dysregulation in chemokine networks (including HGF and CXCL9) that could make them more susceptible to severe COVID-19. Our study also suggest that IL-6 levels are increased in some asthmatics and this may influence their immune response to COVID-19.

## Introduction

During the COVID-19 pandemic, populations at risk of severe symptoms have been detected including patients with asthma and COPD [1]. An analysis of comorbidities in 1,590 COVID-19 patients across China found that COPD patients had a higher risk for intense care unit (ICU) admission, mechanical ventilation, or death, even after adjustment for age and smoking [2]. A recent meta-analysis has reported that COPD is associated with a significant, over fivefold, risk of severe COVID-19 [3]. The risk has been attributed to the overexpression of angiotensin converting enzyme 2 (ACE-2) receptor in bronchial epithelial cells from COPD patients [4] and by the increased expression of the transmembrane protease serine 2 (TMPRSS2) induced by the exposure to cigarette smoking [5]. A recent study also shows that patients with chronic lung disease (including COPD) have baseline changes in cell-type specific expression of genes related to viral replication and the immune response, that could promote immune exhaustion and altered inflammatory gene expression [6], supporting the increased risk in COPD patients. However, the mechanisms remain to be elucidated. On the other hand, it has been found that patients with asthma have increased risk of severe COVID-19 [7]. Also, that severe asthma is associated with COVID-19 related death [8] and asthmatic patients show increased expression of the viral activator TMPRSS2 [9]. Although there is still controversy on whether asthmatics are more susceptible to SARS-CoV-2 infection [10], it is being recognized that endotype, exposome or the presence of other comorbidities may increase risk to severe COVID-19 [11, 12].

As part of our research to identify biological markers for asthma and COPD, we conformed in 2018 the “Identification of Biomarkers in Asthma and COPD cohort (IBACO)” in two pneumology centres in Colombia and collected plasma for biomarker discovery aiming to identify biomarkers that can reveal mechanistical pathways in these diseases, especially to define endotypes of asthma and COPD. Plasma profiling is a useful tool that can reflect systemic inflammatory status including lung tissues [13-16]. We here implemented the multiplexed measurement of 92 plasma proteins based on the proximity ligation assay to provide a comprehensive overview of protein levels in the same sample at the same time [17, 18].

Likewise, there is a growing interest in identifying protein biomarkers of severe COVID-19 that may allow to profile high risk patients for early management and to define point-of-care clinical classifiers [19-21]. Several cohorts worldwide have reported significant changes in protein biomarkers in patients with COVID-19. Initial studies reported molecules that are indicative of inflammation (such as C reactive protein) or changes in cell proportions (neutrophil-to-lymphocyte ratio, NLR) [22, 23], but more recent studies are focused on elucidating pathways underlying patient severity with detailed profiles of plasma molecules such as cytokines, chemokines and growth factors[21, 24-26]. For instance, Messner *et al*., identified 27 proteins in the blood of COVID-19 patients that were present at different levels depending on the severity of their symptoms including complement factors, the coagulation system, inflammation modulators, and pro-inflammatory factors upstream and downstream interleukin 6 [27]. In addition, Arunachalam *et al*., identified a linear increase in the protein levels of MCP-3, TNF, EN-RAGE and TNFSF14, being highest in patients with severe disease and ICU admission [24]. During the analyses of the plasma profiling in the participants of the IBACO cohort, we found that COPD patients show increased levels of several immune proteins that have been reported as upregulated in patients with severe COVID-19 infection. Here we present these proteins and apply induced network analysis to evaluate their functional relationships, trying to understand how their dysregulation influences the susceptibility to severe COVID-19.

## Materials and Methods

### Study participants

The plasma samples were obtained from the “Identification of Biomarkers in Asthma and COPD cohort (IBACO)”, aimed to identify biomarkers of chronic respiratory diseases in Colombia. Patients were recruited from two reference pneumology clinics of Cartagena and Bogotá. The study was approved by the ethical committees of the University of Cartagena (nr. 4169722017) and the “Fundación Neumológica Colombiana” (nr. 232-07122017) and written informed consent was obtained from all participants. The study included a well characterized group of adult subjects aged 40 to 90 years with asthma (n=118) or COPD (n=99) recruited between February 2018 and March 2020, before the first case of SARS-Cov2 infection was reported in the country. Healthy controls (n=99) were recruited during the same period in elderly homes and in the communities with a similar age and gender distribution. At the time of sampling all subjects were queried about their current and past sociodemographic characteristics, symptoms, comorbidities, smoking habits, environmental exposures, history of allergies, pharmacological treatments, and received a physical examination and pulmonary function tests. The diagnosis was done by a pulmonologist according to the GEMA guidelines for asthma [28] and to the GesEPOC guidelines for COPD [29]. Pulmonary function test was done by spirometry pre- and post-bronchodilator according to the American Thoracic Society (ATS) guidelines. Quality of life was assessed by the asthma control questionnaire (ACQ-5), the COPD Assessment Test (CAT), and the Saint George’s Respiratory Questionnaire (SGRQ). Comorbidities were evaluated by the Charlson comorbidity index [30]. Inclusion criteria were no exacerbation in the last eight weeks, age 40 or more, with a clinical diagnosis of asthma or COPD confirmed by a pneumologist and signed informed consent. For COPD patients additional inclusion criteria were a reported exposure to wood smoke for at least 10 years and/or cigarette smoking with more than 10 packs/year. Exclusion criteria included exacerbation of asthma or COPD in the last 8 weeks, presence of uncontrolled comorbidities such as hypertension, coronary disease, hepatic and/or renal diseases, active neoplastic disease, treatment with immunosuppressive drugs, human immunodeficiency virus (HIV) infection, report of respiratory or non-respiratory infection in the last 8 weeks and/or being under treatment with monoclonal antibodies.

### Sample collection

Blood samples were collected by standard phlebotomy in heparinized tubes and plasma was separated by centrifuging at 1000 *g* at 4°C for 15 minutes and stored at −80°C until analysis. Another sample was collected in an EDTA tube to measure leukocyte cell counts by type IV hemocytometry. IgE antibodies were measured by ImmunoCAP following manufacturer instructions (Thermo Fisher, Uppsala, Sweden).

### Quantification of plasma proteins

For plasma profiling the samples were randomly distributed in 96-well plates and protein levels were measured by the Proximity Extension Assay (PEA) [18] using the Target 96 Inflammation Panel (Olink Proteomics, Analysis Service Facility, Boston, USA) which includes a broad selection of proteins established as inflammatory signatures in diverse inflammatory diseases. A total of 67 out of 92 plasma molecules were detected in heparinized plasma (73%). Normalized Protein Levels were expressed as NPX units (log2 scale). Intraassay and interassay average coefficient of variations (%CV) were 6% and 11%, respectively. Nine samples were removed because they did not pass the quality control (QC). Twenty-five proteins had values below the limit of detection (LOD) and were removed from analysis.

### Statistical Analysis

Differences between protein levels among the study groups (asthma, COPD and healthy controls) were first screened by the F-test (ANOVA) using the Olink Insights Stat Analysis Web tool (https://olinkproteomics.shinyapps.io/OlinkInsightsStatAnalysis/) and by the non-parametric Kruskal Wallis test. Given that some of the proteins with significant differences between groups did not have a normal distribution (Kolmogorov-Smirnov test), we implemented both independent samples *t*-test and Mann-Witney tests for comparing protein levels between patients and controls. Correlation coefficients were calculated by the Pearson test. A *P* ≤ 0.05 was considered significant. To adjust for multiple testing, the Benjamini–Hochberg false discovery rate (FDR) correction was applied using the p.adjust function. Statistical analyses were performed in R version 3.5.3 (https://www.r-project.org/).

### Functional annotation

The list of proteins with differences between patients and healthy controls were analysed for pathways, ontologies, diseases/drugs in the Enrichr web tool (https://maayanlab.cloud/Enrichr/) [31]. Protein connections and pathways were analysed in the PathwAX web tool (https://pathwax.sbc.su.se/help.html) [32]. The induced network analysis was constructed in Consensus PathDB (http://cpdb.molgen.mpg.de/) using an intermediate nodes z-score threshold of 30 and including binary protein-protein interactions of high confidence and biochemical reactions [33].

### Biomarkers of severe COVID-19

We performed a literature search on all proteins described associated with severe COVID-19. We started with a set of 93831 articles downloaded on January 30th, 2021 from LitCovid, a curated open-source literature of Pubmed research papers related to COVID-19. Then we retrieved a corpus of documents on severe COVID-19 using the terms (“severe COVID” or “fatal COVID”) AND (“biomarker” OR “plasma marker” OR “plasma protein”). This resulted in 114 papers of which 20 reported associations of severe COVID-19 with 52 unique proteins. The protein list from this study was also compared to evaluate the enrichment with that reported in the COVID-19 Related Gene Sets (SARS 133 Literature-Associated Genes from Geneshot GeneRIF) and calculate the probability of detecting overlap due to chance.

## Results

Demographic and clinical characteristics of the study participants are presented in **Table 1**. As expected, asthmatic patients have higher IgE levels and eosinophils than healthy controls and COPD patients. On the other hand, COPD patients have higher numbers of neutrophils and monocytes while there were no differences in the number of lymphocytes. A total of 315 plasma samples were analysed with the Olink Inflammation panel to compare proteins levels between groups (**Figure 1A**).

**Table 1.** Demographic and clinical characteristics of study subjects.

| Variables | Healthy controls<br>(n=98) | Asthma patients<br>(n=118) | COPD patients<br>(n=99) | P value |
| --- | --- | --- | --- | --- |
| Age, yr | 56 ± 13 | 60 ± 11 | 72 ± 8 | <0.0001 |
| Female gender, n (%) | 63 (64.3%) | 82 (69.5%) | 40 (40.4%) | <0.0001 |
| BMI, kg/m<sup>2</sup> | 24.9 ± 4.5 | 27.5 ± 4.9 | 25.1 ± 4.5 | <0.0001 |
| Tabaquisism ever, yes, n (%) | 22 (22.4%) | 40 (33.9%) | 91 (91.9%) | <0.0001 |
| Pack/year, n | 1.1 ± 4.0 | 1.72 ± 5.6 | 32.1 ± 26.7 | <0.0001 |
| Passive smoking, yes, n (%) | 33 (33.7%) | 56 (47.5%) | 51 (51.5%) | 0.03 |
| Exposure to wood smoke, yes, n (%) | 17 (17.3%) | 26 (22%) | 32 (32.3%) | 0.04 |
| Age of wheezing onset, n (%) |  |  |  | - |
| <5 | n/a | 7 (5.9%) | 0 (0%) |  |
| 5-14 | n/a | 33 (28.0%) | 1 (53.5%) |  |
| 15-40 | n/a | 38 (32.2%) | 8 (52.5%) |  |
| >40 | n/a | 32 (27.1%) | 44 (44.4%) |  |
| ER visits in the last year due to exacerbation of respiratory symptoms | 0 | 1.2 ± 3 | 0.6 ± 1.1 | <0.0001 |
| mMRC | 0.2 ± 0.5 | 1.1 ± 0.9 | 1.6 ± 1.0 | <0.0001 |
| ACQ-5 | n/a | 4.54 ± 4.4 | n/a | - |
| CAT | n/a | n/a | 13.8 ± 7.3 | - |
| Pre-FEV<sub>1</sub> % | 96.3 ± 18.4 | 71.1 ± 20.5 | 55.3 ± 20.1 | <0.0001 |
| Post BD FEV<sub>1</sub> % | 98 ± 18.1 | 77.2 ± 21.9 | 59 ± 21.1 | <0.0001 |
| Pre FEV<sub>1</sub>/FVC, % | 80 ± 6 | 69 ± 12 | 60 ± 13 | <0.0001 |
| Post BD FEV<sub>1</sub>/FVC, % | 82 ± 6 | 71 ± 12 | 60 ± 13 | <0.0001 |
| Blood eosinophils, cells/μl | 140 (90-192) | 220 (120-312) | 190 (130-310) | <0.0001 |
| Blood neutrophils, cells/μl | 3090 (2517-3862) | 3815 (3122-4940) | 4170 (3430-5310) | <0.0001 |
| Blood monocytes, cells/μl | 435 (357-525) | 500 (390-602) | 540 (450-680) | <0.0001 |
| Blood lymphocytes, cells/ μl | 2185 (1720-2600) | 2145 (1670-2642) | 1950 (1590-2440) | 0.13 |
| Total, IgE, kU/l | 57.7 (21-189.7) | 134.3 (44.4-370) | 71.9 (23.6-175.4) | < 0.0001 |
| IgE to *D. pteronyssinus* | 0.04 (0.03-0.21) | 0.24 (0.04-6.54) | 0.05(0.03-0.24) | < 0.0001 |
| IgE to *B. tropicalis* | 0.02 (0.00-0.11) | 0.21 (0.01-2.39) | 0.02 (0.00-0.35) | < 0.0001 |
| IgE to *A. lumbricoides* | 0.02 (0.00-0.07) | 0.05 (0.01-0.36) | 0.03 (0.00-0.15) | < 0.0001 |
| Inhaled corticosteroid dose |  |  |  | - |
| High | 0 (0%) | 34 (28.8%) | 2 (2%) |  |
| Medium | 0 (0%) | 35 (29.7%) | 19 (19.2%) |  |
| Low | 0 (0%) | 42 (35.6) | 5 (5.1%) |  |
| None | 0 (0%) | 5 (4.2%) | 68 (68.7%) |  |
| Charlson comorbidity index | 0.05±0.26 | 1.2 ± 0.6 | 1.3 ± 0.7 | <0.0001 |
| Cardiovascular disease, yes, n (%) | 0 (0%) | 5 (4.2%) | 11 (11.1%) | - |
| Diabetes, yes, n (%) | 1 (1%) | 8 (6.8%) | 11 (11.1%) | - |

BMI: Body Mass Index, ER: emergency room, mMRC: modified Medical Research Council dyspnea scale; ACQ-5: asthma control questionnaire; CAT: COPD Assessment Test, FEV<sub>1</sub>: forced expiratory volume in 1 second; FVC: forced vital capacity.

**Figure 1.**
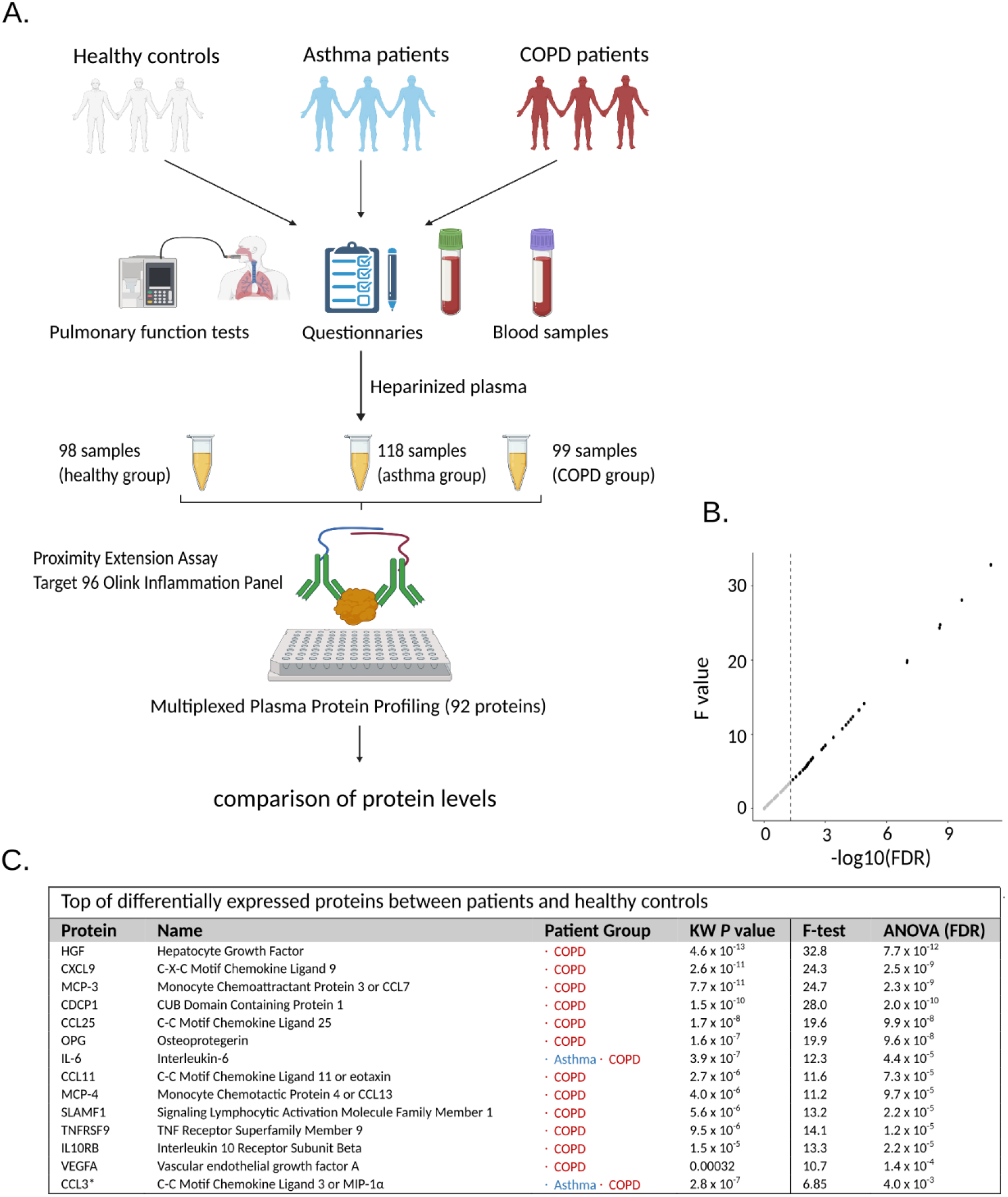
A) Schematic representation of study groups and samples included in the proteomic screening. B) Dot plot on the F value versus the False Discovery Rate (FDR) for all measured proteins. Dotted lines represent *P* = 0.05. C) Top differentially expressed proteins by comparing asthma and COPD patients with healthy controls. Detailed information on protein names and *P* values is presented in Supplementary table 1.

### Patients with asthma and COPD have increased levels of several plasma inflammatory proteins

We found differences in 41 plasma proteins between patients and healthy controls (Kruskal Wallis test, nominal P < 0.05). Similar results were obtained by the analysis of variance (ANOVA) and 36 proteins remained significant after Benjamini–Hochberg false discovery rate (FDR) correction **(Figure 1B)**. The top significant proteins are presented in **Figure 1C**.

We then compared protein levels between each disease group and healthy controls by t-test. The levels of 8 proteins showed differences in asthmatic patients (IL-6, CXCL1, MMP-1, CSF-1, CXCL5, CCL3, CCL23 and TNFSF14, nominal P < 0.05) while 40 proteins showed significant differences in COPD patients. However, when applying the FDR correction for multiple testing, only IL-6 remained significant in asthma patients (**Figure 2A**) and 39 proteins remained significant in COPD patients (**Figure 2B**).

**Figure 2.**
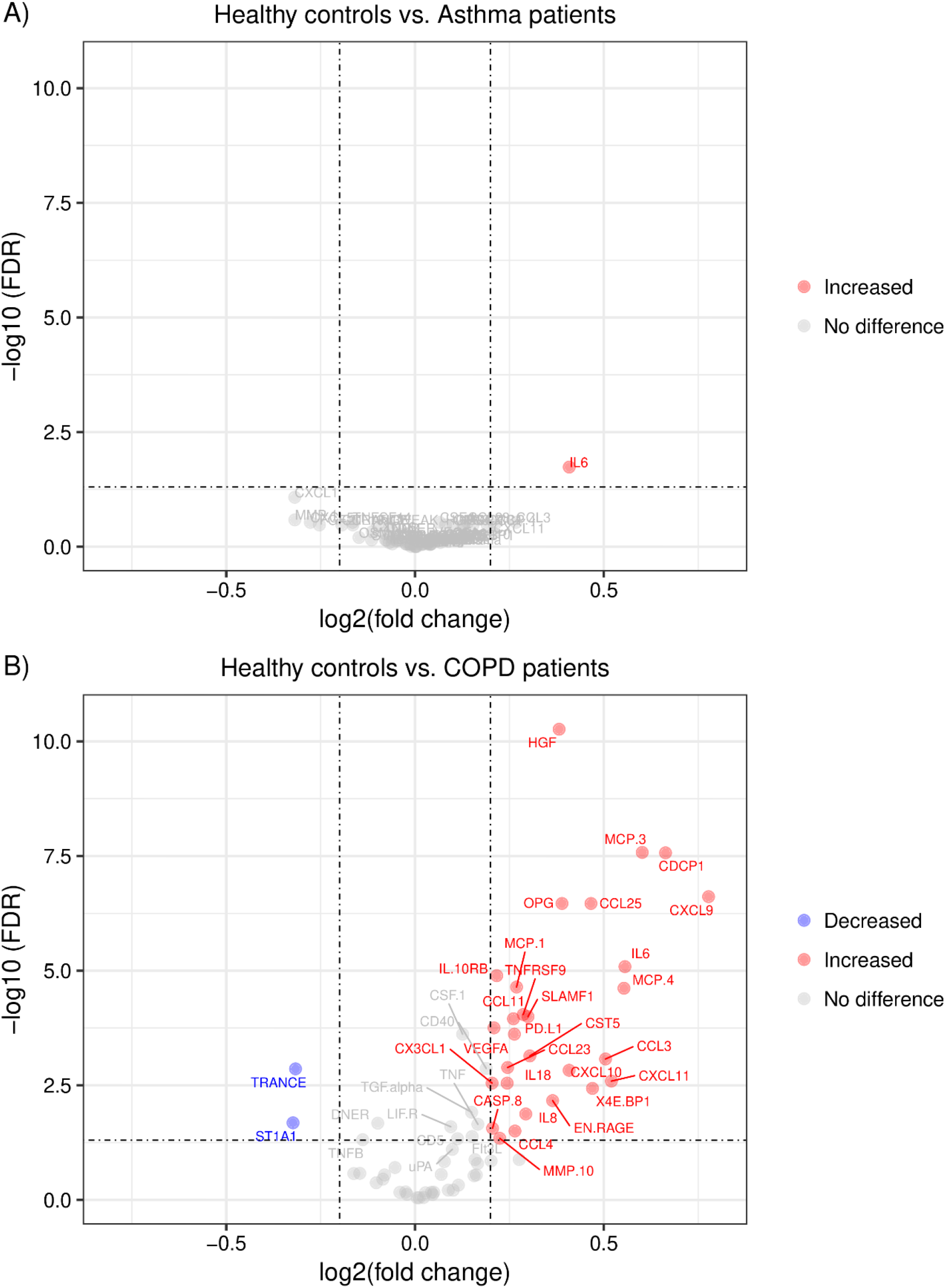
Volcano plots of differentially expressed plasma proteins between patients and healthy controls. Lines indicate cut-off points for significance (FDR < 0.05 and fold change ≥0.2).

### Differentially expressed proteins are grouped in different clusters

We also performed correlation analysis to evaluate the relationships between proteins levels and identify co-regulated signatures in 30 proteins with significant differences in the COPD patients (FDR < 0.05 and fold change > 0.20). We found two correlation clusters: the first within seven proteins (OPG, HGF, CXC3L1, VEGFA, IL10RB, TNFRSF9, PD-L1) exhibiting high to moderate correlation. The strongest coefficients were observed between TNFRSF9 and VEGFA (r = 0.73, *P* = 1.1 × 10^−17^), TNFRSF9 and IL10RB (r = 0.73, *P* = 9.9x 10^−18^) and HGF and OPG (r = 0.72, *P* = 8.6 × 10^−17^).

The second cluster involved eight chemokines with moderate correlation in their protein levels (CCL11, MCP-1, MCP-3, CXCL11, MCP-4, IL-8, CXCL9 and CXCL10). The most significant were CXCL11 and MCP-4 (r = 0.67, *P* = 1.9 × 10^−14^), CXCL9 and CXCL10 (r = 0.65, *P* = 5.1 × 10^−13^), MCP-3 and MCP-4 (r = 0.58, *P* = 2.1 × 10^−10^) and CXCL11 and CXCL10 (r = 0.57, *P* = 5.3 × 10^−10^). These analyses also showed that CXCL9 levels correlate with proteins in the first cluster and TRANCE (RANKL) had an inverse correlation with its ligand OPG (r = −0.27, *P* = 5.5 × 10^−3^) (**Figure 3**).

**Figure 3.**
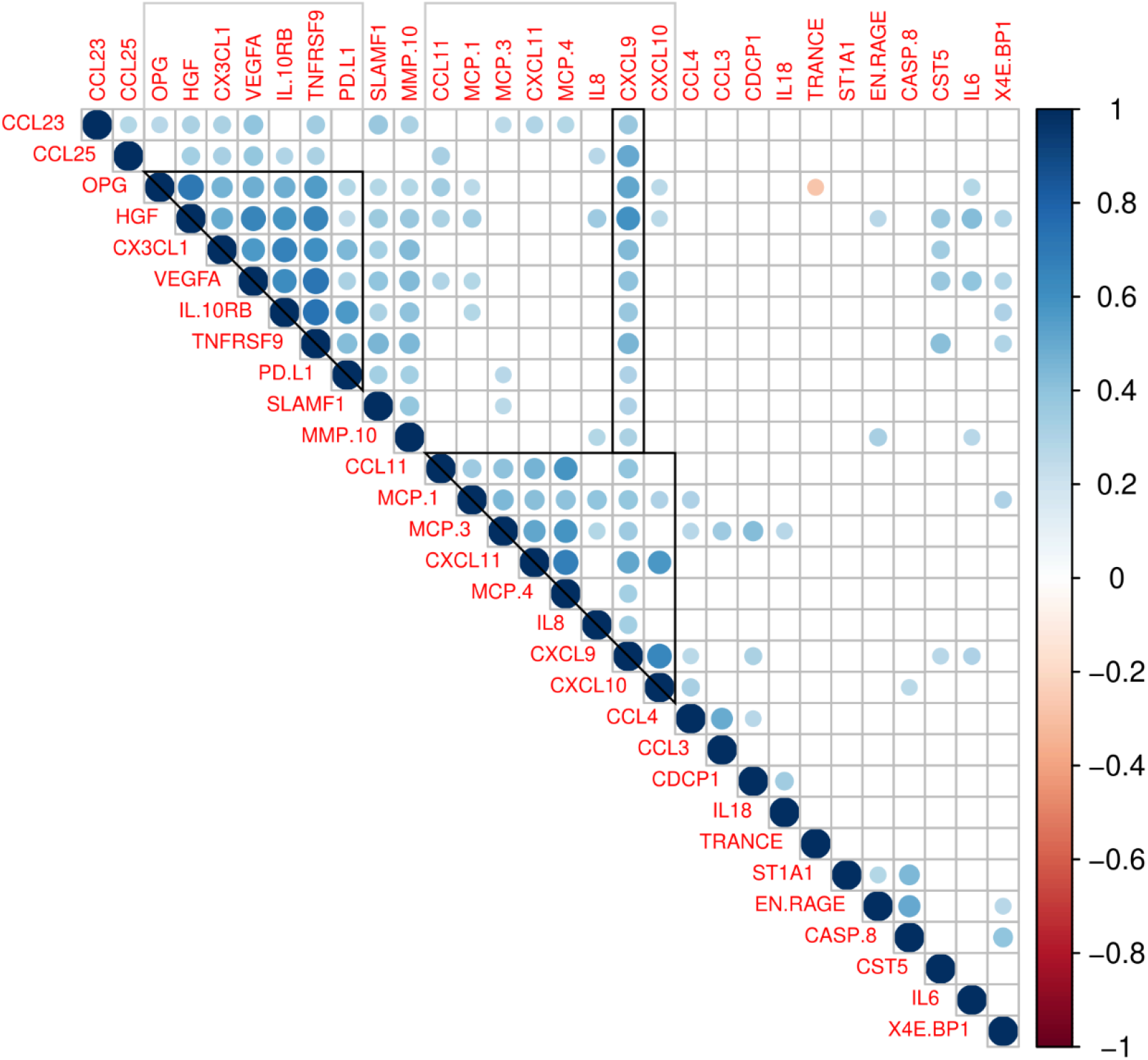
Correlations between plasma levels in 30 plasma proteins showing significant differences between COPD patients and controls with FDR < 0.05 and fold change > 0.20. Coloured circles indicate a significant correlation (P < 0.05). The colour scale represents the Pearson correlation coefficient (1 positive correlation: −1 negative correlation). Grey squares indicate clusters of correlated proteins.

### Differentially expressed proteins are involved in recognized inflammatory pathways

To further evaluate the biological relationships between the correlated protein clusters we performed enrichment analyses on pathways and gene ontology terms. Differentially expressed proteins were enriched in the Gene Ontology (GO) categories of cytokine and chemokine signaling pathways (**Figure 4A**). The analysis on PathwAX showed the top significant enrichment in the pathways of “Viral protein interaction with cytokine and cytokine receptor”, “TNF signaling pathway” and the “NF-kappa B signaling pathway” (**Figure 4B**). The proteins that overlap with these biological processes involve several chemokines.

**Figure 4.**
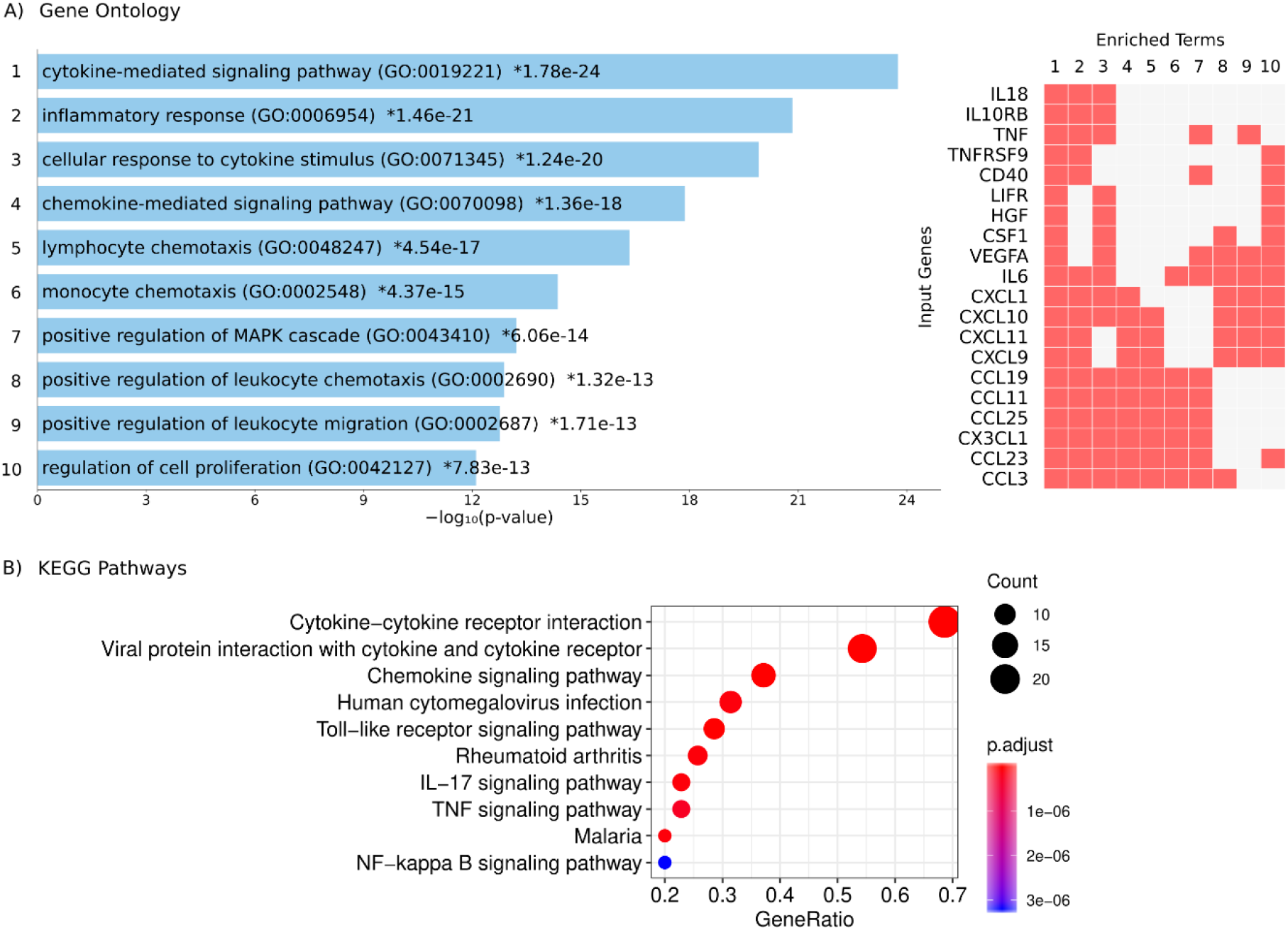
Functional annotation of the differentially expressed plasma proteins. A). Gene Ontology analysis: each bar represents a significant GO term with its *P* value in the enrichment analysis. Asterisks indicate significant after correction for multiple testing. The proteins detected in this study that coincide with those involved in each biological process (1 to 10) are indicated by a red box (right panel). B). Significant pathways on the Kyoto Encyclopedia of Genes and Genomes (KEGG).

In addition, we explored the biological connections among differentially expressed proteins using them as seeds in an induced network analysis considering evidence of binary protein-protein interactions at high level of confidence and biochemical reactions. We found several interactions between the cluster of HGF, OPG, TRANCE and VEGFA with the cluster of chemokines. First, the strong correlation of HGF with OPG could be explained with their interaction mediated by the von Villebrand factor (VWF). Upon that OPG interacts with its ligand TRANCE, metalloproteinases (MMP) and the chemokine MCP-3. Besides, an interaction mediated by heparan sulphate (HS) links HGF with IL-8 and VEGFA. The interaction of HGF with the chemokine cluster is also mediated by Platelet Factor 4 (PF4). HGF also interacts with TNF involving neuropilin 1 (NRP1) **(Figure 5)**.

**Figure 5.**
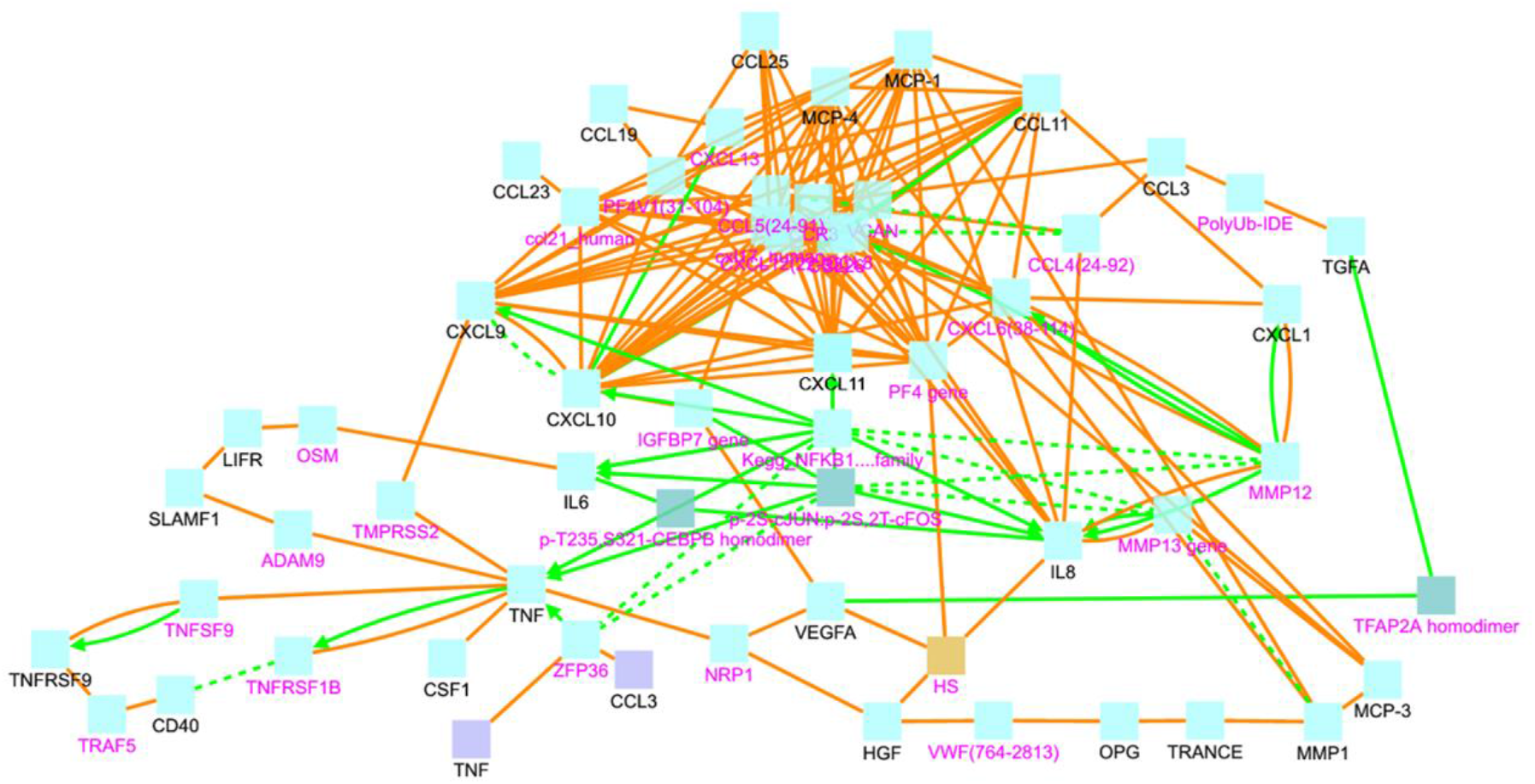
Induced network analysis showing the protein-protein interactions (orange line) and biochemical reactions (green lines) among the differentially expressed proteins (black letters) or mediated by interconnecting nodes (magenta letters). VWF: von Villebrand Factor. HS: heparan sulphate.

We also observed an interaction between TNF and CXCL9 that has TMPRSS2 as an intermediate node. From there, CXCL9 and CXCL10 seem to be immune checkpoints connected with several other chemokines. IL-6 and IL-8 are interconnected between them and with the chemokine cluster by members of the NFĸB signaling pathway **(Figure 5)**.

### Asthma, COPD and severe COVID-19 patients shared increased plasma inflammatory proteins

The differentially expressed proteins were not only involved in biological pathways of viral immune response (**Figure 4**) but were also found enriched in datasets of genes upregulated by SARS coronaviruses. The COVID-19 Drug and Gene Set Library retrieved at least 10 experiments with significant enrichment of the genes encoding the altered proteins detected in this study upon infection. The top significant association was with nine genes upregulated by SARS-CoV-2 in Calu3 cells including CXCL9, TNFRSF9, CX3CL1, CSF1, LIFR, IL6, CXCL11, CXCL10 and TNF (**Figure 6)**.

**Figure 6.**
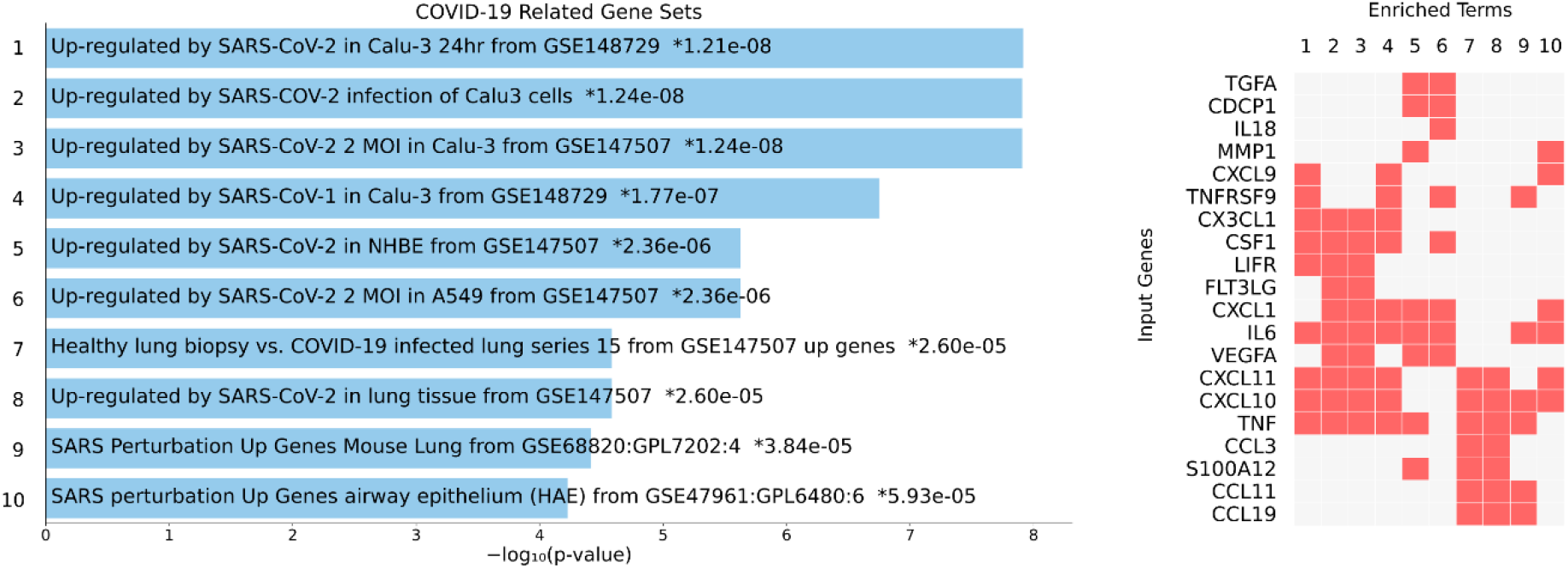
Overlap between genes upregulated upon coronavirus infection with the proteins detected in this study. The details on the experiment are indicated on the blue bars (left) with the *P* value for the enrichment analysis. Asterisks indicate significant enrichment after correction for multiple testing. The proteins detected in this study that coincide with the genes found in each experiment (1 to 10) are indicated by a red box (right panel).

A comparison with 133 SARS Literature-Associated Genes in Geneshot GeneRIF revealed significant overlap with four proteins found increased in our patients (CXCL10, CXCL9, IL-6, TNF, *P* = 6.9 × 10^−11^). We then analysed which of the differentially expressed proteins detected in this study have been previously reported associated with the severity of SARS-Cov-2 infection in humans. Strikingly, we found that the top significant protein HGF have been reported increased in critically ill COVID-19 patients [25, 34]. Also, other proteins such as IL-6, MCP-3, CXCL10, TNF, and EN-RAGE have been found increased in patients with severe COVID-19 [24]. The protein levels of eight severe COVID-19 biomarkers (HGF, CXCL9, IL-8, IL-6, MCP-3, VEGFA, CXCL10, CCL3) as detected in healthy controls, asthmatics and COPD patients from this study are presented in **Figure 7**.

**Figure 7.**
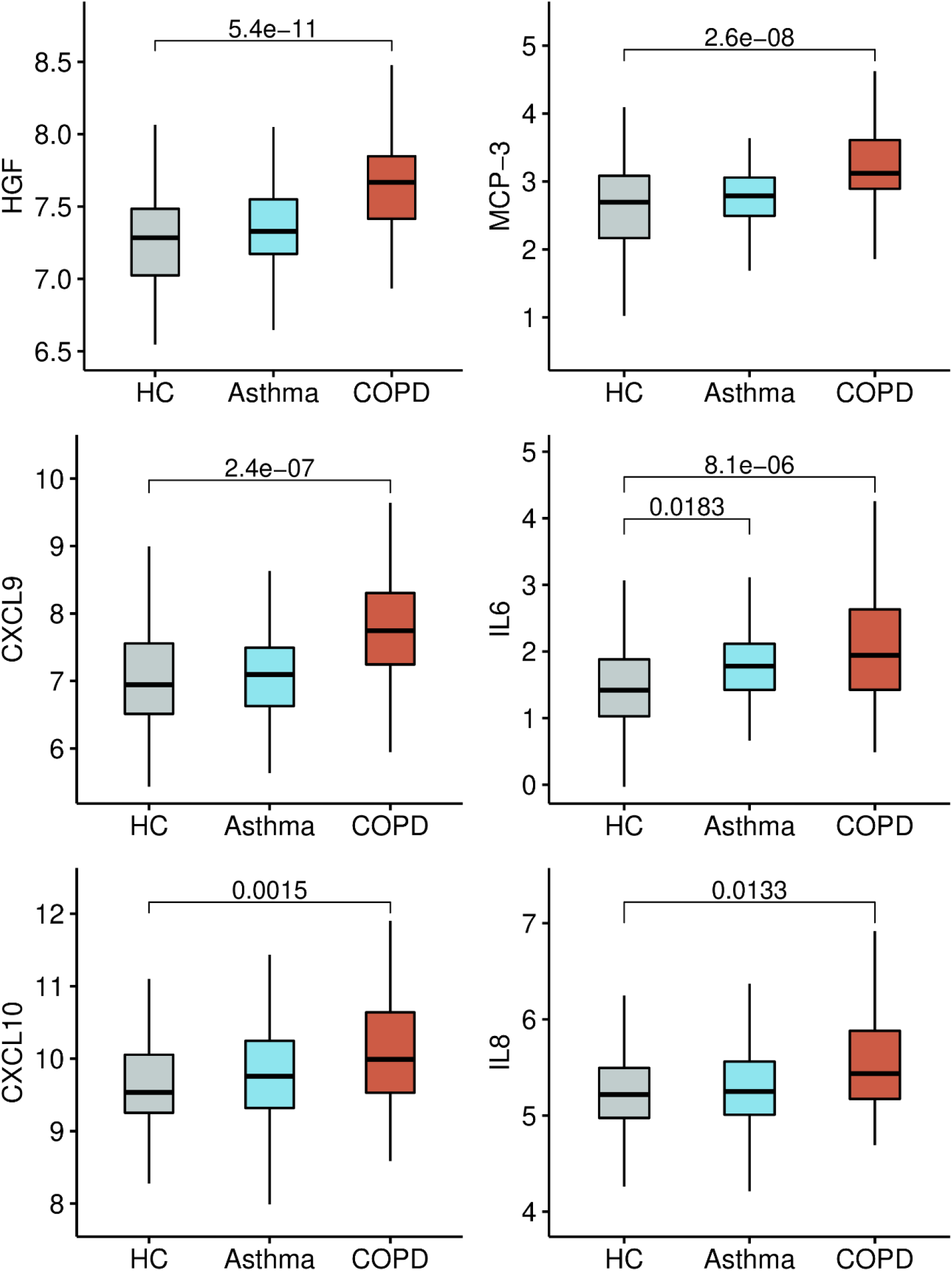
Boxplots with the normalized protein levels (log2 scale) of eight proteins associated with severe COVID-19 in non-infected healthy controls (HC), asthma and COPD patients.

### Several plasma proteins associated with severe COVID-19 are influenced by age

Plasma levels in 18 proteins showed significant correlations with age. The highest coefficients were detected with OPG (r=0.59, *P* = 9.3 × 10^−11^), HGF (r=0.48, *P* = 3.2 × 10^−7^) and CXCL9 (r=0.43, *P* =7.6 × 10^−6^). Of these age-correlated proteins, six had been also associated with COVID-19 severity: HGF and CXCL9 shown the highest coefficients while IL10RB, VEGFA, IL-6 and TNF showed low albeit significant correlation (r < 0.30). Besides, MCP-3, EN-RAGE and IL-8 did not show relation with age. The dynamics of nine biomarkers of severe COVID-19 are presented in **Figure 8**.

**Figure 8.**
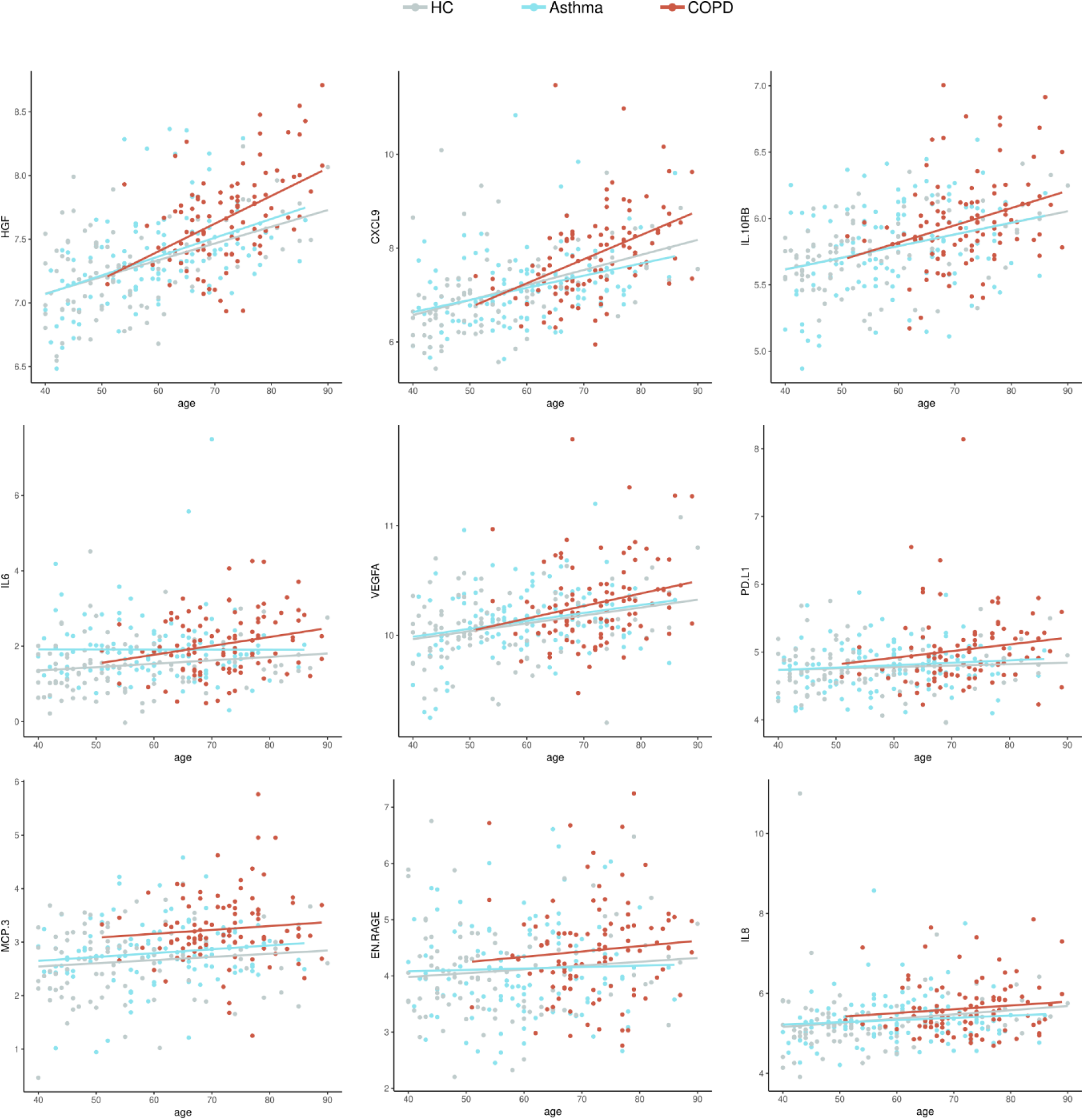
Dot plots with the normalized protein levels (log2 scale) in relation to age, each dot represent a subject. Lines represent the regression.

## Discussion

This study revealed that patients with COPD have significant differences in several plasma proteins compared to healthy controls. Among the proteins increased in COPD patients, HGF, CXCL9, MCP-3, IL-6, IL-8, CXL10, EN-RAGE, IL10RB, VEGFA, CCL3 and TNF have been also found upregulated in patients with severe COVID-19 [24, 26] and some of them such as HGF postulated as biomarkers for the severe form of this infection [21, 34, 35]. The overlap between these biomarkers of COVID-19 severity and the proteins we found increased in plasma from COPD patients add important information regarding the susceptibility of these patients to severe COVID-19. Asthmatic patients only showed increased levels in IL-6 compared to healthy controls (**Figure 2A**), suggesting that a more systemic inflammation in COPD patients might contribute to their increased risk for severe COVID-19.

HGF was the top significant protein marker in our study. Deng *et al*, reported that HGF levels above 1,128 pg/ml can discriminate severe from non-severe COVID-19 patients, with an 84.6% sensitivity and 97.9% specificity to classify severe patients (AUC of 90.5%). Also, they report that HGF might significantly increase only when inflammation mounts toward an uncontrolled storm [34]. However, our results demonstrate that patients with COPD have a significant elevation of this marker before getting infected with SARS-CoV-2; therefore, further studies are needed to elucidate the dynamic and importance of HGF in COPD and COVID-19 patients.

Additionally, plasma levels of HGF, osteoprotegerin (OPG) and TRANCE were correlated and change in aged healthy groups. Given the relationship of OPG and TRANCE with osteoclastogenesis [36, 37], our data may reflect protein changes due to aging, but also indicate that the HGF-OPG-TRANCE axis is altered at earlier age-points in COPD patients (**Figure 9**). The relevance of this finding on the increased susceptibility of aged individuals to severe COVID-19 needs to be further defined.

**Figure 9.**
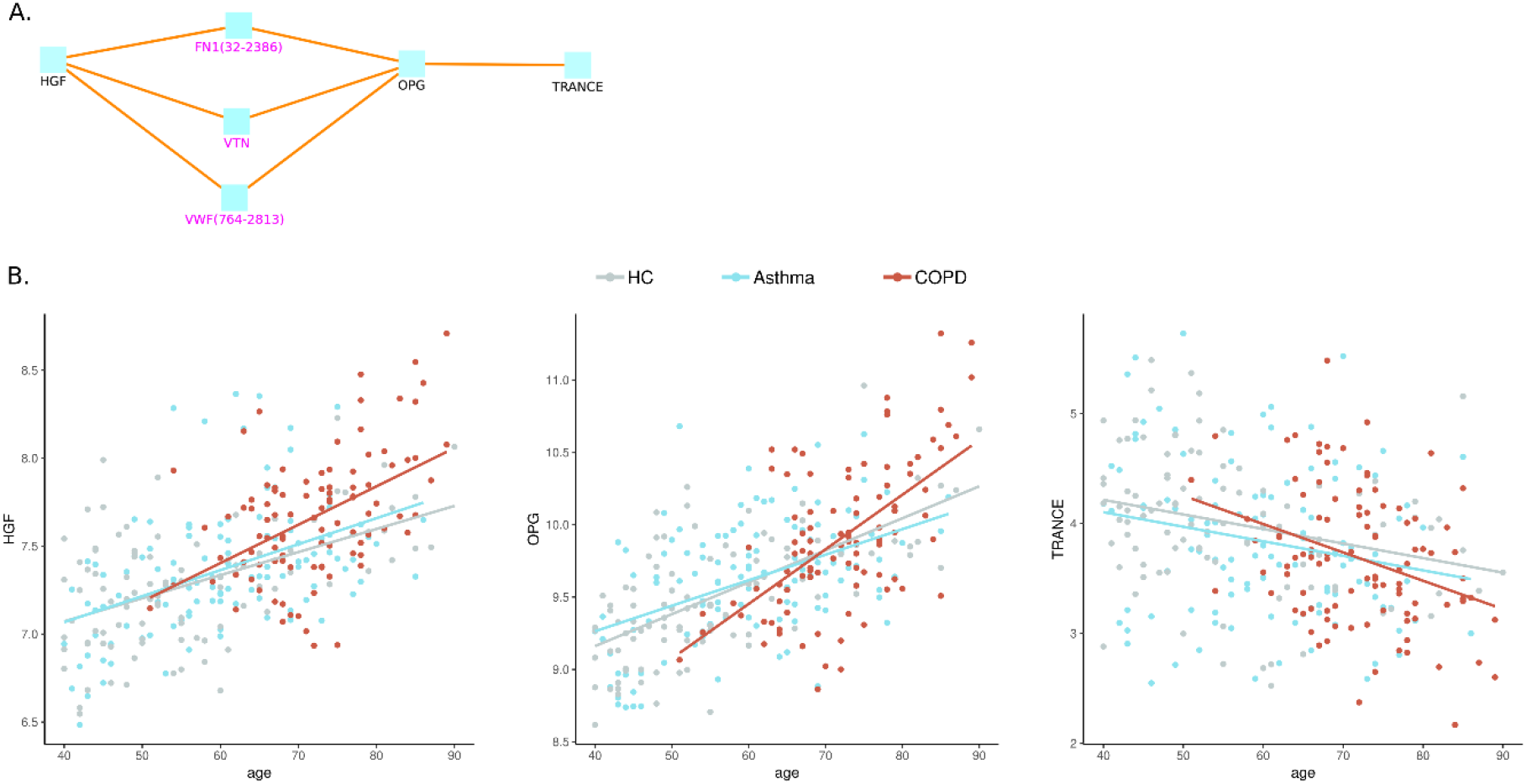
The HGF, OPG and TRANCE axis in patients and controls. A) Schema on the protein-protein interactions between HGF and OPG mediated by fibronectin (FN1), vitronectin (VTN) and the von-Willebrand factor (VWF). B) Dot plots with the normalized protein levels (log2 scale) in relation to age, each dot represent a subject. Lines represent the regression.

Other studies have linked HGF with a neutrophil signature that predicts critically ill COVID-19 patients [38]. In that study, HGF levels directly correlate with absolute neutrophil counts (rho=0.55, *P* < 0.001) and together with resistin and lipocalin-2 are released from neutrophil granules upon activation. Indeed, HGF and three other neutrophil granule proteins (RETN, LCN2, MMP-8) were not elevated in non-ICU patients compared to controls and were only significantly different in patients who would go on to develop critical illness [38]. The COPD patients in this study have increased levels of HGF and higher numbers of neutrophils compared to asthmatics and healthy controls, however we did not find correlation between HGF and neutrophil counts in COPD patients (data not shown).

Our induced network analysis revealed protein-protein interactions between HGF and OPG mediated by the von Willebrand factor (VWF) suggesting a connection of these markers with the coagulation system (**Figure 5**). In the study of Thwaites *et al*., the von-Willebrand factor A2 was elevated in all hospitalized COVID-19 patients and the elevation was highest in severely affected COVID-19 patients [39]. Other studies also support the elevation of VWF as risk factor for severe COVID-19 [40]. In this setting, our data support that HGF and OPG could be involved in severe COVID-19 by affecting pathways implicated in coagulopathy and thrombosis [41-43]. Since HGF have been reported increased in patients with rheumatic diseases [44] as well as in patients with insulin resistance-diabetes [45], obesity [46], and diabetic kidney disease [47], further studies are needed to establish if it is elevated in severe COVID-19 as a pre-existing condition reflecting the presence of the comorbidity, or if increases during the infection and has a role in the pathophysiology this condition.

Other cytokines such as IL-6 and TNF-α have been documented in severe COVID-19 [48]. Indeed, high serum IL-6, IL-8 and TNF-α levels at the time of hospitalization were strong and independent predictors of patient survival [20]. Here we found increased levels of IL-6, IL-8 and TNF in COPD patients. The analysis of protein-protein interactions between the differentially expressed proteins detected in this study, revealed an interaction between TNF and CXCL9 that has the viral activator TMPRSS2 as intermediate node (**Figure 5**). This transmembrane serine protease has been found increased in smokers and in COPD patients [5, 49], and mediates the cleavage of the S protein in SARS-CoV-2, thus is instrumental for viral entry [50]. CXCL9 was the protein showing the second most significant differences between COPD patients and healthy controls (**Figure 1 and 2**). The consequences of these protein interactions need to be experimentally elucidated, however our results suggest that altered levels of TNF, IL-6 and CXCL9 in COPD patients may be involved in the initial steps that predispose COPD patients to severe COVID-19. CXCL9 is a C-X-C motif chemokine induced by interferon, considered a marker of monocyte/macrophage activation that is implicated in lymphocyte infiltration, Th1 polarization and antiviral immune responses [51].Several studies have reported increased expression of CXCL9 in severe COVID-19 patients [52-55], as well as in CXCL10 and CXCL11 [24, 56], three co-expressed chemokines that were found increased in the COPD patients in our study **(Figure 2 and 7)**.

Another interesting finding was the enrichment of increased proteins in COPD patients in chemokine and cytokine networks associated to viral response **(Figure 4)**. Respiratory viral infections can trigger acute exacerbations in COPD patients. Different viruses are implicated, for example rhinovirus, influenza, syncytial respiratory virus and coronavirus [57]; being rhinovirus and coronavirus detected in 35.7% and 25.9% of all viral infections occurring during acute exacerbations [58]. Typically, COPD patients are more susceptible to viral infections. The mechanisms underlying this susceptibility included dysregulated antiviral function of CD8^+^ T cells via the PD-1/PD-L1 axis [59] and an altered production of interferons and chemokines [60]. Thus, it is possible that differences in protein plasma levels and the enrichment of differentially expressed proteins in viral related pathways **(Figure 4)**, could be the consequence of previous viral infections that induced lung inflammation and altered the plasma proteome profile in our COPD patients.

However, since our COPD patients had not experienced exacerbations in at least eight weeks before sampling, these differences could reflect sub-clinical viral infections as has been described in patients with stable COPD [61, 62]. Changes in some of these proteins and especially in chemokines, might reflect an altered immune response that could explain their increased susceptibility to a broad spectrum of viral infections including SARS-CoV-2.

In conclusion, our results provide evidence of dysregulated inflammatory signatures in the plasma proteome of COPD that may underlie their increased susceptibility to severe COVID-19. Since these proteins were found increased in patients without SARS-CoV-2 infection, they could be further evaluated as biomarkers of increased risk to severe COVID-19. The detection of dysregulated plasma proteins in other conditions known to be highly susceptible to severe COVID-19 will provide important information for preventing and treating this viral disease. Moreover, the proteins detected in this study provide potential targets of new or repositioned therapeutic approaches aiming to avoid severe COVID-19 in patients with COPD.

## Data Availability

All the data regarding this manuscript is available from the corresponding author on request

## Acknowledgments

We thank Mileidis Avila, Ana Carolina Mercado, Ivan Acevedo Monterrosa, Tania Cepeda and Patricia Parada for their technical support during patient recruitment and sample processing. To Dina Ortiz for her skilful assistance with the pulmonary function tests and to all the patients and healthy participants that voluntarily contributed to this study.

## Abbreviations

CCL3: C-C motif chemokine ligand 3 or Macrophage inflammatory protein 1-alpha
CCL4: C-C motif chemokine ligand 4 or Macrophage inflammatory protein 1-beta
CCL11: C-C Motif chemokine ligand 11 or eotaxin-1
COPD: Chronic obstructive pulmonary disease
COVID-19: Coronavirus disease 19
CSF-1: Colony stimulating factor 1
CST5: Cystatin-D
CX3CL1: C-X3-C motif chemokine ligand 1 (fractalkine)
CXCL1: C-X-C motif chemokine ligand 1 or Neutrophil-activating protein 3
CXCL5: C-X-C motif chemokine ligand 5 or Neutrophil-activating protein 78
CXCL9: C-X-C motif chemokine ligand 9 or Monokine induced by interferon-gamma
CXCL10: C-X-C motif chemokine ligand 10 or IP-10
CXCL11: C-X-C motif chemokine ligand 11 or interferon-inducible T-cell alpha chemoattractant
EN-RAGE: extracellular newly identified receptor for advanced glycation end-products binding protein or S100 Calcium Binding Protein A12
HGF: Hepatocyte Growth Factor or lung fibroblast-derived mitogen
IL10RB: Interleukin 10 receptor subunit beta
KEGG: Kyoto Encyclopedia of Genes and Genomes
MCP-3: Monocyte chemoattractant protein 3 or CCL7
MCP-4: Monocyte chemotactic protein 4 or CCL13
MMP-1: Matrix metallopeptidase 1
OPG: Osteoprotegerin or Tumor necrosis factor receptor superfamily, member 11b
SARS-CoV-2: Severe acute respiratory syndrome coronavirus 2
SLAF1: Signaling lymphocytic activation molecule family member 1
ST1A1: Sulfotransferase family 1A member 1
TMPRSS2: Transmembrane serine protease 2
TNF: Tumor Necrosis Factor
TNFRSF9: TNF receptor superfamily member 9
TNFSF14: TNF superfamily member 14
TRANCE: TNF superfamily member 11 or Receptor activator of Nuclear Factor Kappa B Ligand
VEGFA: Vascular Endothelial Growth Factor A

---

**Supplementary table 1.**
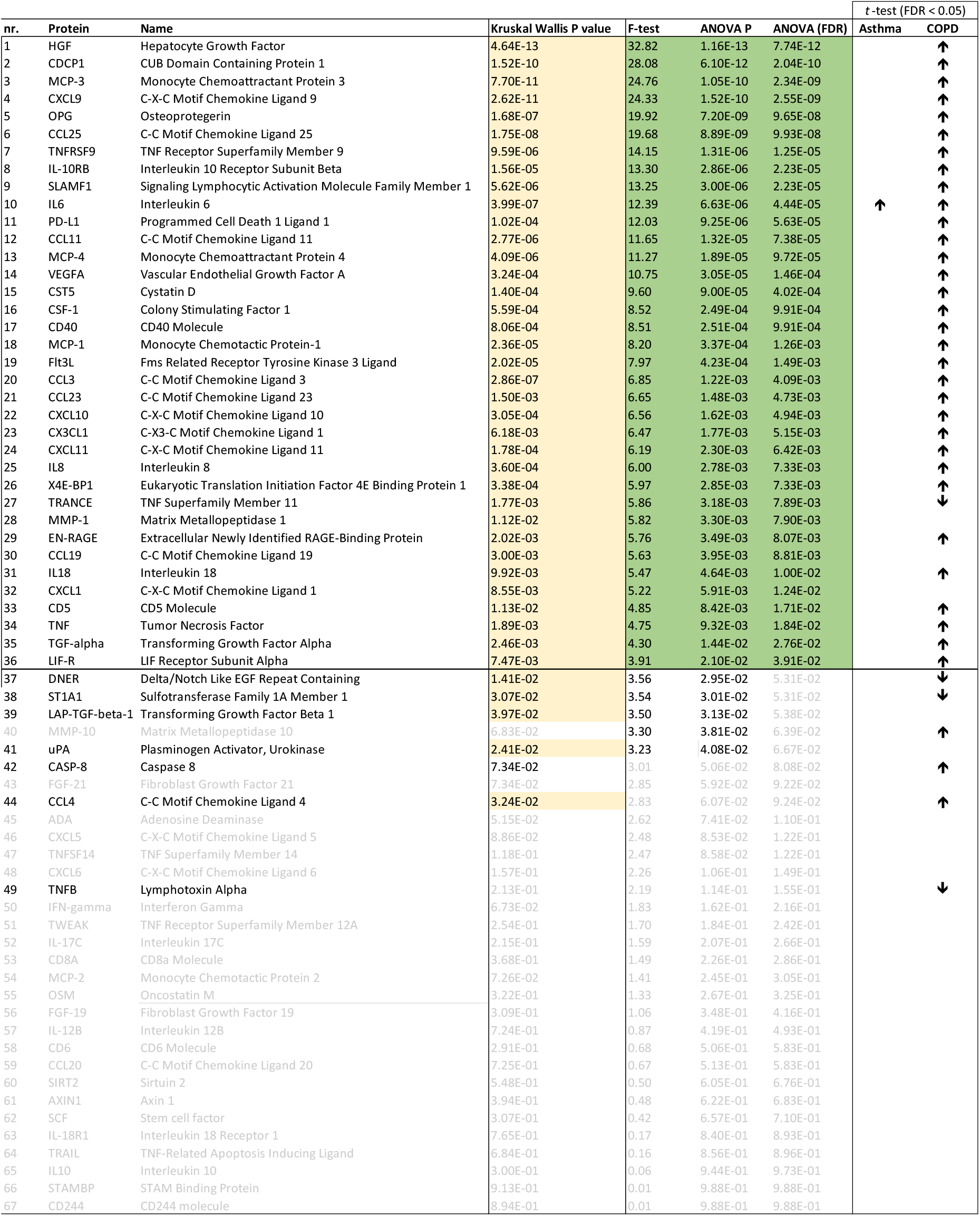
Annotation of the differentially expressed proteins between patients and healthy controls

## Notes

### Competing Interest Statement

The authors have declared no competing interest.

### Clinical Trial

This is not a clinical trial.

### Funding Statement

This study was financed by the Ministry of Science (Republic of Colombia, Grant 756-2017) and the University of Cartagena

### Author Declarations

The plasma samples were obtained from the Identification of Biomarkers in Asthma and COPD cohort (IBACO), aimed to identify biomarkers of chronic respiratory diseases in Colombia. Patients were recruited from two reference pneumology clinics of Cartagena and Bogota. The study was approved by the ethical committees of the University of Cartagena (nr. 4169722017) and the Fundacion Neumologica Colombiana (nr. 232-07122017) and written informed consent was obtained from all participants.

### Summary of Updates

The figures has been revised to add FDR p values and improved resolution.

